# High-sensitivity cardiac troponin on presentation to rule out myocardial infarction: a stepped-wedge cluster randomised controlled trial

**DOI:** 10.1101/2020.09.06.20189308

**Authors:** Atul Anand, Kuan Ken Lee, Andrew R. Chapman, Amy V. Ferry, Phil D. Adamson, Fiona E. Strachan, Colin Berry, Iain Findlay, Anne Cruikshank, Alan Reid, Paul O. Collinson, Fred S. Apple, David A. McAllister, Donogh Maguire, Keith A.A. Fox, David E. Newby, Chris Tuck, Ronald Harkess, Catriona Keerie, Christopher J. Weir, Richard A. Parker, Alasdair Gray, Anoop S.V. Shah, Nicholas L. Mills, on behalf of the HiSTORIC Investigators

**Affiliations:** BHF Centre for Cardiovascular Science, University of Edinburgh, Edinburgh, UK; Christchurch Heart Institute, University of Otago, Christchurch, New Zealand; Institute of Cardiovascular and Medical Sciences, University of Glasgow, Glasgow, UK; Department of Cardiology, Royal Alexandra Hospital, Paisley, UK; Department of Biochemistry, Queen Elizabeth University Hospital, Glasgow, UK; Departments of Clinical Blood Sciences and Cardiology, St George’s, University Hospitals NHS Trust and St George’s University of London, London, UK; Department of Laboratory Medicine and Pathology, University of Minnesota, Minneapolis, MN, USA; Institute of Health and Wellbeing, University of Glasgow, Glasgow, UK; Emergency Medicine Department, Glasgow Royal Infirmary, Glasgow, UK; Edinburgh Clinical Trials Unit, Usher Institute, University of Edinburgh, Edinburgh, UK; Emergency Medicine Research Group Edinburgh, Royal Infirmary of Edinburgh, Edinburgh, UK; Usher Institute, University of Edinburgh, Edinburgh, UK

**Author notes:** Contributed equally (Atul Anand, Kuan Ken Lee, Andrew R. Chapman, Anoop S.V. Shah, Nicholas L. Mills). Listed in the Supplement. **Corresponding Author:** Professor Nicholas L Mills BHF/University Centre for Cardiovascular Science The University of Edinburgh Edinburgh EH16 4SA United Kingdom.

## Abstract

**Objectives:** High-sensitivity cardiac troponin assays enable myocardial infarction to be ruled out earlier, but the efficacy and safety of this approach is uncertain. We investigated whether an early-rule out pathway is safe and effective for the management of patients with suspected acute coronary syndrome.

**Design:** A stepped-wedge cluster randomised controlled trial.

**Setting:** Seven acute care hospitals in Scotland.

**Participants:** 31,492 consecutive patients with suspected acute coronary syndrome between December 2014 to December 2016.

**Intervention:** Sites were randomised to implement an early rule-out pathway where myocardial infarction was ruled out if high-sensitivity cardiac troponin I concentrations were <5 ng/L at presentation. During a prior validation phase, myocardial infarction was ruled out where troponin concentrations were < 99_th_ centile at 6-12 hours after symptom onset.

**Main outcome measures:** The co-primary outcome was length of stay (efficacy), and myocardial infarction or cardiac death after discharge at 30 days (safety). Patients were followed for 1 year to evaluate the safety outcome and other secondary outcomes.

**Results:** We enrolled 31,492 patients (59±17 years, 45% women) with troponin concentrations < 99_th_ centile at presentation. The length of stay was reduced from 10.1±4.1 to 6.8±3.9 hours (adjusted geometric mean ratio 0.78, 95% confidence interval [CI] 0.73 to 0.83, P< 0.001) following implementation, and the proportion of patients discharged increased from 50% to 71% (adjusted odds ratio [aOR] 1.59, 95% CI 1.45 to 1.75). Non-inferiority was not demonstrated for the 30-day safety outcome (upper limit of one-sided 95% CI for adjusted risk difference 0.70%, non-inferiority margin 0.50%, P = 0.068), but the observed differences favoured the early rule-out pathway (0.4% [57/14,700] *versus* 0.3% [56/16,792]). At 1 year, the safety outcome occurred in 2.7% (396/14,700) and 1.8% (307/16,792) of patients before and after implementation (aOR 1.02, 95% CI 0.74 to 1.40, P = 0.894), and there were no differences in hospital reattendance or all-cause mortality.

**Conclusions:** Implementation of an early rule-out pathway for myocardial infarction reduced length of stay and hospital admission. Whilst non-inferiority for the safety outcome was not demonstrated at 30 days, there was no increase in cardiac events at 1 year. Adoption of this pathway will have major benefits for patients and healthcare providers.

**Trial registration:** http://ClinicalTrials.gov number, NCT03005158

## Introduction

There are over 20 million presentations with suspected acute coronary syndrome each year in the US alone,_1_ accounting for up to a tenth of hospital visits and 40 percent of unscheduled admissions._2_ Given that most patients do not have myocardial infarction,_3_ the adoption of effective and safe pathways to rule out myocardial infarction in the Emergency Department and avoid hospital admission would have a major impact on patient care and healthcare provision.

Cardiac troponin testing is an integral component of the assessment of patients with suspected acute coronary syndrome, with guidelines recommending serial testing at presentation and 6-12 hours later to coincide with the peak in troponin concentration._4_ The development of high-sensitivity cardiac troponin assays with enhanced precision at very low concentrations permits quantification well below the 99_th_ centile diagnostic threshold for myocardial infarction._5_ This advance has led to innovative pathways to rule out myocardial infarction more rapidly, either at presentation or within 3 hours._6–13_ However, these studies were observational, and there are few examples where the pathway guided patient care._14_,_15_ The majority were modest in size, or enrolled selected low-risk patients, and therefore the true efficacy and safety of introducing these pathways into clinical practice remains uncertain.

Our aim was to determine the efficacy and safety of implementing an accelerated pathway where high-sensitivity cardiac troponin testing is used to rule out myocardial infarction at presentation in consecutive patients with suspected acute coronary syndrome.

## Methods

### Trial Design and Oversight

*Hi*gh-*S*ensitivity cardiac *T*roponin *O*n presentation to *R*ule out myocardial *I*nfar*C*tion (*HiSTORIC*) is a stepped-wedge cluster randomised controlled trial enrolling consecutive patients with suspected acute coronary syndrome across seven acute hospitals in Scotland. In this trial, the hospital site was the unit of randomisation. The trial was approved by the Scotland A Research Ethics Committee and the conduct of the trial was periodically reviewed by an independent trial steering committee. All data were collected from the patient record and national registries, deidentified and linked in a data repository (DataLochTM, Edinburgh, UK) within secure NHS safe havens._16_

### Trial Population

Sites were eligible if they had the capacity to introduce the early rule-out pathway and returned data to the national registry. All patients in the Emergency Department were identified by the attending clinician using an electronic form integrated into the care pathway at the time troponin was requested. Patients were eligible for inclusion if they presented with suspected acute coronary syndrome and had a troponin concentration within the normal reference range at presentation. Patients were excluded if they presented with an out-of-hospital cardiac arrest or ST-segment elevation myocardial infarction, had been admitted previously during the trial or were not resident in Scotland.

### Randomisation

The trial was conducted across three phases (***Figure 1a***). During a validation phase of 6-9 months, troponin testing was performed at presentation and repeated 6-12 hours after the onset of symptoms if indicated (standard care). In accordance with guidelines at the time of enrolment,_4_,_17_ myocardial infarction was ruled out where troponin concentrations were less than the 99_th_ centile at presentation if symptom onset was > 6 hours from presentation, or following serial testing 6-12 hours from symptom onset. Sites were paired based on the expected number of patients and randomised to implement the early rule-out pathway (intervention) in one of three steps during a 6-month randomisation phase. Finally, all sites completed an implementation phase of 6-9 months calendar matched to the validation phase where care was guided by the early rule-out pathway.

**Figure 1.**
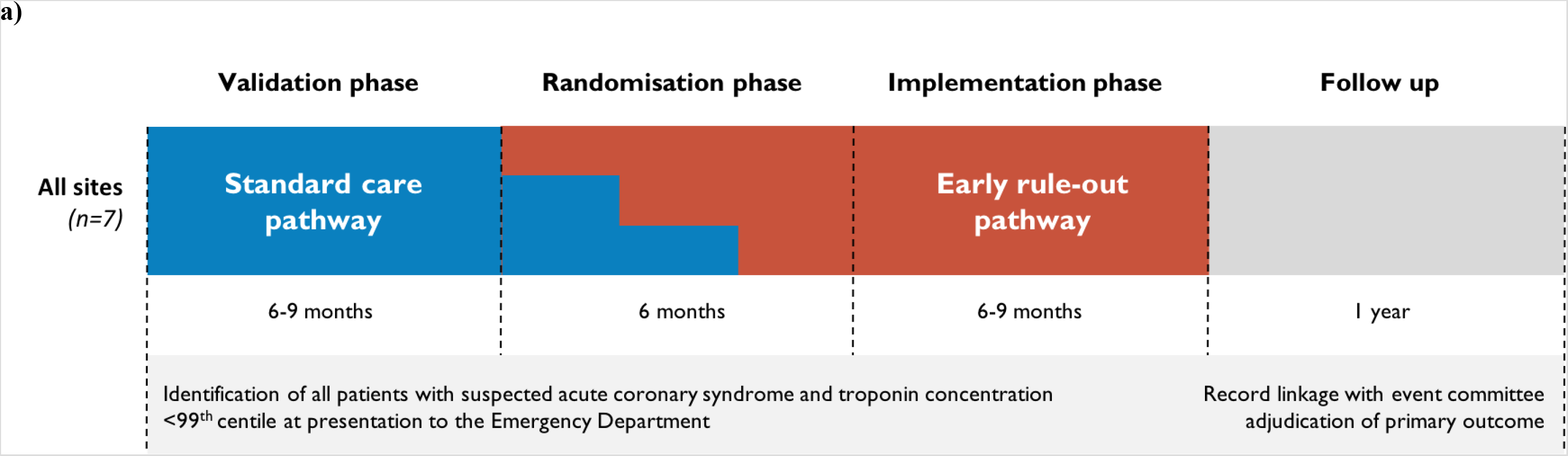
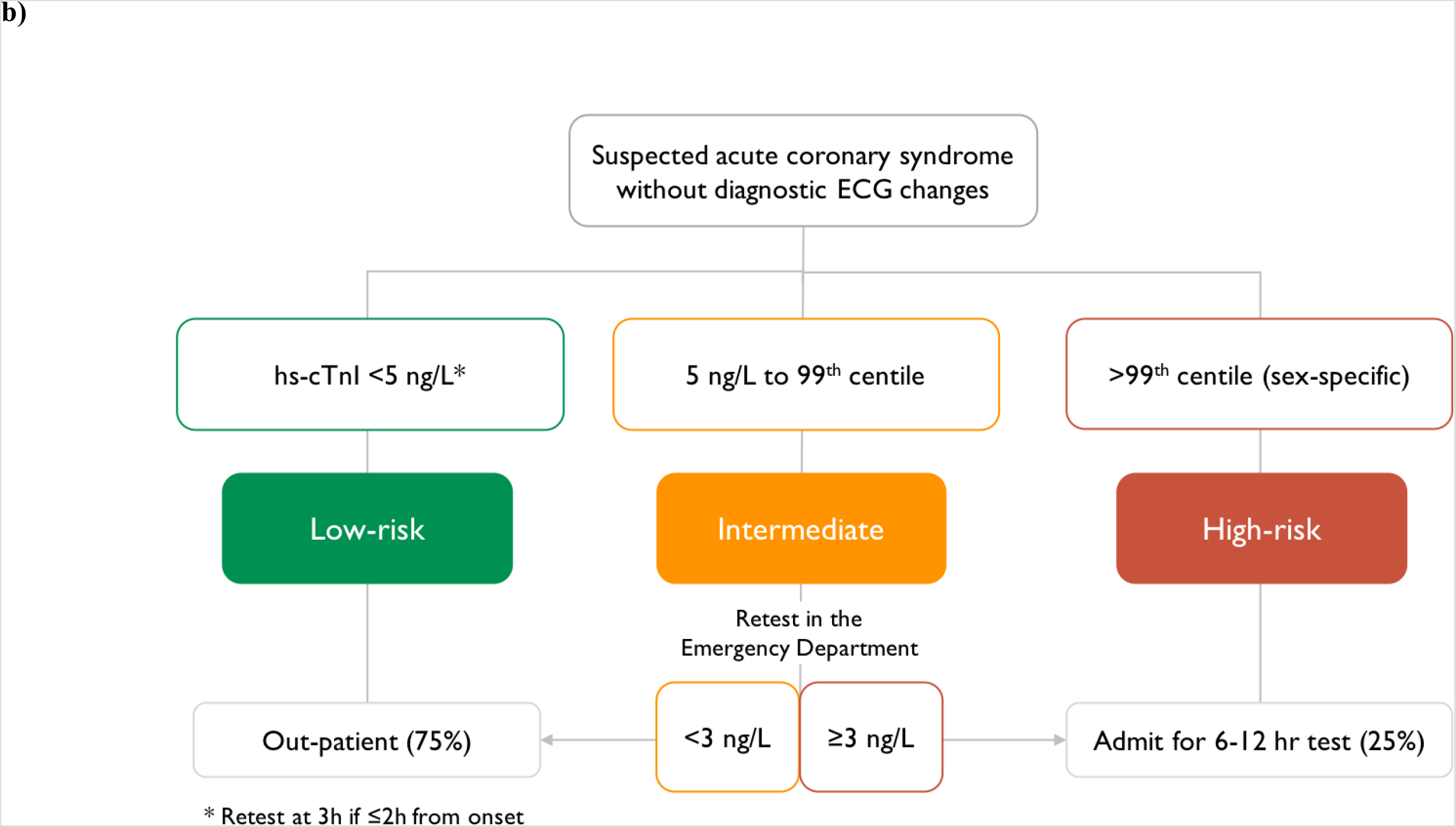
Schematic diagram of the HiSTORIC trial design and the early-rule out pathway. **a)** During a validation phase of at least 6 months, cardiac troponin testing was performed at presentation and was repeated 6 or 12 hours after the onset of symptoms with myocardial infarction ruled out where cardiac troponin concentrations were less than the sex-specific 99_th_ centile (standard care). Sites were paired based on the expected number of patients and randomised to implement the early rule out pathway pathway (intervention) in one of three steps during a 6 month randomisation phase. Finally, all sites completed an implementation phase of at least 6 months that was calendar matched to the validation phase where patient care was guided by the early rule-out pathway. **b)** The early rule-out pathway rules out myocardial infarction at presentation in patients with cardiac troponin concentrations below a risk stratification threshold of 5 ng/L, unless they presented within 2 hours of symptom onset where testing is repeated 3 hours from presentation. Patients with cardiac troponin concentrations ≥5 ng/L at presentation are retested in the Emergency Department 3 hours after presentation, and myocardial infarction is ruled out if concentrations are unchanged (delta < 3 ng/L) and remain below the 99_th_ centile diagnostic threshold.

### Intervention

The *High*-*S*ensitivity *T*roponin in the *E*valuation of patients with suspected *A*cute *C*oronary *S*yndrome (*High-STEACS*) early rule-out pathway (***Figure 1b***) has been described previously._18_,_19_ Myocardial infarction is ruled out in patients with troponin concentrations < 5 ng/L at presentation, unless they present within 2 hours of symptom onset where testing is repeated 3 hours from presentation. Patients with troponin concentrations ≥5 ng/L at presentation are retested 3 hours after presentation, and myocardial infarction is ruled out if concentrations are unchanged (delta < 3 ng/L) and remain below the 99_th_ centile. To support implementation, we provided educational material and presentations at each site, a webapp (http://www.highsteacs.com), and formal training for clinical staff in the Emergency Department (***eMethods in Supplement***). Throughout the trial, all sites used the Abbott ARCHITECT*STAT* high-sensitive troponin I assay to guide clinical decisions. This assay has an inter-assay coefficient of variation of less than 10% at 4.7 ng/L,_8_,_20_ and a 99_th_ centile of 16 ng/L in women and 34 ng/L in men._21_

### Trial Outcomes

We used regional and national registries to follow-up the trial population._16_,_22-23_ Sequential hypothesis testing evaluated two co-primary outcomes in an *a-priori* defined hierarchical order: the primary efficacy outcome followed by the primary safety outcome. The primary efficacy outcome was length of stay, defined as the length of time from presentation to the Emergency Department until discharge from hospital. The safety outcome was myocardial infarction (type 1, type 4b or type 4c) or cardiac death after discharge which was evaluated at 30 days (primary) and 1 year (secondary) following presentation. These events were adjudicated by a panel blind to the study phase. All subsequent presentations where any troponin concentration was > 99_th_ centile were reviewed and adjudicated as described previously (***eMethods in Supplement***)._16_,_24_,_25_

The secondary efficacy outcome measure was the proportion of patients discharged from the Emergency Department. Other safety outcome measures included myocardial infarction, cardiac death, cardiovascular death, all-cause death, unplanned coronary revascularisation and reattendances for any reason after discharge at 1 year. Adherence was evaluated for three prespecified components of the early rule-out pathway (***eMethods in Supplement***).

### Statistical Analysis

All outcomes were analysed using a linear mixed-effects regression model, adjusting for hospital site (random effect), season, time of presentation since start of study, and an indicator variable for whether the early rule-out pathway had been implemented. Length of stay was log-transformed prior to analysis and results expressed as a geometric mean ratio. If the analysis of the primary efficacy outcome was significant at the 5% level, then we planned to perform a non-inferiority analysis of the primary safety outcome reporting a risk difference (intervention–standard care) and one-sided 95% confidence interval. If the upper limit of the one-sided 95% confidence interval was below a 0.5% non-inferiority margin, then non-inferiority was established, and if it was below 0% then superiority was established. A sensitivity analysis compared outcomes during the calendar matched period in the validation and implementation phases using the same regression model as for the primary analysis, but without adjustment for time or season. A number of other sensitivity analyses were performed (***eMethods in Supplement***).

### Patient and public involvement

A patient review panel was consulted throughout the trial programme and provided input on the educational advice provided to clinicians following introduction of the new pathway. Qualitative research capturing the views and experiences of patients treated within these pathways will follow in a separate publication. Patients were not involved in the conception or design of the trial.

## Results

### Trial Sites and Population

Seven acute hospitals were eligible and all participated (***Table S1***). Between December, 2014, and December, 2016, a total of 31,492 consecutive patients with suspected acute coronary syndrome (59±17 years, 45% women) met the inclusion criteria (***Figure S1***). There were 14,700 (47%) and 16,792 (53%) patients assessed before and after implementation of the early rule-out pathway, respectively. Clinical characteristics were similar before and after implementation (***Table 1***) and across all three phases of the trial (***Table S1***). The trial concluded in December, 2017 with 1 year of follow up available in 31,428 (99.8%) patients.

**Table 1.** Characteristics of the Trial Participants.

|  | All | Standard care | Early rule-out |
| --- | --- | --- | --- |
| No. of participants | 31,492 | 14,700 | 16,792 |
| Age (years) | 59±17 | 59±17 | 58±17 |
| Women | 14,252 (45) | 6,575 (45) | 7,677 (46) |
| ***Presenting complaint*** |  |  |  |
| Chest pain | 26,590 (84) | 12,566 (85) | 14,024 (84) |
| Dyspnoea | 957 (3) | 420 (3) | 537 (3) |
| Palpitation | 928 (3) | 432 (3) | 496 (3) |
| Syncope | 1,701 (5) | 699 (5) | 1,002 (6) |
| Other | 1,316 (4) | 583 (4) | 733 (4) |
| ***Past medical history*** |  |  |  |
| Myocardial infarction | 2,573 (8) | 1,371 (9) | 1,202 (7) |
| Ischaemic heart disease | 7,346 (23) | 3,834 (26) | 3,512 (21) |
| Cerebrovascular disease | 1,684 (5) | 849 (6) | 835 (5) |
| Diabetes mellitus | 1,912 (6) | 1,002 (7) | 910 (5) |
| ***Previous revascularisation*** |  |  |  |
| PCI | 2,831 (9) | 1,534 (10) | 1,297 (8) |
| CABG | 452 (1) | 240 (2) | 212 (1) |
| ***Medications at presentation*** |  |  |  |
| Aspirin | 8,023 (25) | 4,114 (28) | 3,909 (23) |
| Dual anti-platelet therapy* | 1,269 (4) | 738 (5) | 531 (3) |
| Statin | 12,165 (39) | 6,035 (41) | 6,130 (37) |
| ACE inhibitor or ARB | 9,769 (31) | 4,776 (32) | 4,993 (30) |
| Beta-blocker | 8,548 (27) | 4,162 (28) | 4,386 (26) |
| Oral anti-coagulant † | 2,167 (7) | 1,033 (7) | 1,134 (7) |
| ***Electrocardiogram ‡*** |  |  |  |
| Normal | 12,035 (74) | 6,118 (73) | 5,917 (75) |
| Myocardial ischemia | 3,288 (20) | 1,756 (21) | 1,532 (20) |
| ST-segment elevation | 193 (1) | 111 (1) | 82 (1) |
| ST-segment depression | 252 (2) | 146 (2) | 106 (1) |
| T-wave inversion | 1,225 (8) | 621 (7) | 604 (8) |
| Other | 1,711 (11) | 927 (11) | 784 (10) |
| ***Haematology and clinical chemistry*** |  |  |  |
| Haemoglobin, g/L | 137±22 | 137±20 | 137±23 |
| eGFR, mL/min | 81±22 | 81±23 | 82±22 |
| Presentation hs-cTnI, ng/L | 3 [1-6] | 3 [1-6] | 3 [1-6] |
| Peak hs-cTnI, ng/L | 3 [1-7] | 3 [1-7] | 3 [1-7] |
| Serial (≥2) tests § | 11,904 (38) | 6,540 (44) | 5,364 (32) |
| ***Time intervals*** |  |  |  |
| Symptom onset to presentation ≤2 hrs | 5,664 (18) | 2,859 (19) | 2,805 (17) |
| Presentation to first test, mins | 66 [45-97] | 66 [46-97] | 65 [43-97] |
| First to second test, mins | 351 [188-553] | 455 [267-601] | 229 [155-405] |

Presented as No. (%), mean±SD or median [inter-quartile range]. Abbreviations: ACE = angiotensin converting enzyme; ARB = angiotensin receptor blockers; eGFR = estimated glomerular filtration rate; CABG = coronary artery bypass grafting; PCI = percutaneous coronary intervention.
\* Two medications from aspirin, clopidogrel, prasugrel or ticagrelor. † Includes warfarin or novel oral anti-coagulants. ‡ Proportions reported for the 16,217 (51%) participants with electrocardiographic data available. § Serial testing was defined as two or more tests within 24 hours of presentation.

### Primary and Secondary Efficacy Outcomes

Length of stay was reduced from 10.1±4.1 to 6.8±3.9 hours (adjusted geometric mean ratio 0.78, 95% confidence interval [CI] 0.73 to 0.83, P< 0.001) following implementation of the early rule-out pathway (***Table 2, Figure 2***). The proportion of patients discharged from the Emergency Department without hospital admission increased from 50% to 71% (adjusted odds ratio 1.59, 95% CI 1.45 to 1.75). Adherence to all three prespecified components of the early rule-out pathway was observed in 11,600/16,792 (69%) of patients.

**Table 2.** Efficacy and Safety Outcomes at 30 Days and 1 Year.

|  | All | Standard care | Early rule-out | Adjusted Odds Ratio (95% CI)* | P-value |
| --- | --- | --- | --- | --- | --- |
| No. of participants | n=31,492 | n=14,700 | n=16,792 |  |  |
| ***Efficacy outcome*** |  |  |  |  |  |
| Length of stay, hrs (primary) | 8.2±4.1 | 10.1±4.1 | 6.8±4.1 | 0.78 (95% CI 0.73 to 0.83) | P<0.001 |
| Discharge from the ED (secondary) | 19,249 (61) | 7,407 (50) | 11,842 (71) | 1.59 (95% CI 1.45 to 1.75) | P<0.001 |
| ***Safety outcome †*** |  |  |  |  |  |
| 30 days (primary) | 113 (0.4) | 57 (0.4) | 56 (0.3) | 1.97 (95% CI 0.95 to 4.08) | P=0.068 |
| 1 year (secondary) | 703 (2.2) | 396 (2.7) | 307 (1.8) | 1.02 (95% CI 0.74 to 1.40) | P=0.894 |
| ***Other safety outcomes at 1 year*** |  |  |  |  |  |
| Myocardial infarction ‡ | 422 (1.3) | 238 (1.6) | 184 (1.1) | 1.10 (95% CI 0.72 to 1.68) | P=0.646 |
| Cardiac death | 319 (1.0) | 176 (1.2) | 143 (0.9) | 1.07 (95% CI 0.69 to 1.64) | P=0.771 |
| Cardiovascular death | 452 (1.4) | 249 (1.7) | 203 (1.2) | 0.93 (95% CI 0.66 to 1.32) | P=0.696 |
| All-cause death | 1,720 (5.5) | 852 (5.8) | 868 (5.2) | 0.92 (95% CI 0.75 to 1.12) | P=0.385 |
| Unplanned revascularisation § | 222 (0.7) | 119 (0.8) | 103 (0.6) | 0.60 (95% CI 0.35 to 1.03) | P=0.065 |
| Any hospital reattendance | 12,306 (39.1) | 5,770 (39.3) | 6,536 (38.9) | 0.93 (95% CI 0.84 to 1.02) | P=0.112 |

Presented as geometric mean ± standard deviation or No. (%). Abbreviations: CI = confidence interval, ED = Emergency Department
\* Outcomes following implementation of the early rule-out pathway are compared to those during standard care for all measures.
† Type 1, type 4b or type 4c myocardial infarction or cardiac death following discharge.
‡ Type 1, type 4b or type 4c myocardial infarction
§ Unplanned revascularisation was defined as urgent or emergency percutaneous coronary intervention or coronary artery bypass grafting from discharge to 1 year

**Figure 2.**
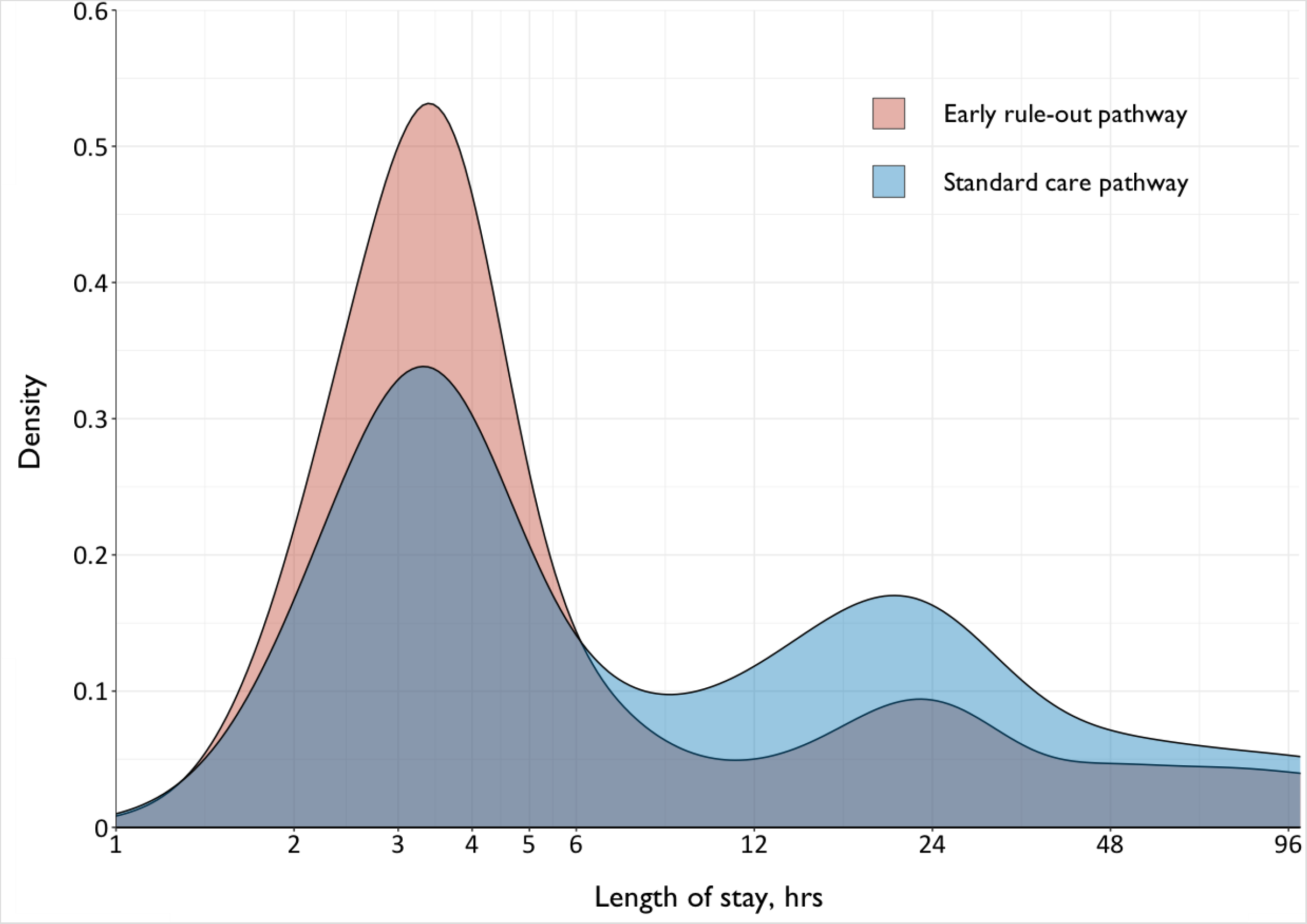
Length of stay before and after implementation of the early rule-out pathway. Shown is a density plot of the length of stay in patients evaluated before (blue) and after (red) implementation of the early rule-out pathway.

### Primary and Secondary Safety Outcomes

Before and after implementation of the early rule-out pathway, the primary safety outcome of myocardial infarction or cardiac death following discharge at 30 days occurred in 57/14,700 (0.4%) and 56/16,792 (0.3%) patients respectively (***Table 2**)* with an adjusted odds ratio of 1.97 (95% CI 0.95 to 4.08, P = 0.068). Comparing the rate of the primary safety outcome after implementation to the rate before, the upper limit of our one-sided 95% confidence interval for the adjusted risk difference was 0.70%, exceeding our prespecified non-inferiority margin of 0.50%. The event rate at 30 days was lower than anticipated, and our regression model and prespecified sensitivity analyses gave divergent results (***Table S3***). However, there were 703 (2.2%) patients with myocardial infarction or cardiac death following discharge at 1 year (***Figure 3***). Before and after implementation, the secondary safety outcome measure occurred in 396/14,700 (2.7%) and 307/16,792 (1.8%) patients, respectively (adjusted odds ratio 1.02, 95% CI 0.74 to 1.40, P = 0.894). Furthermore, the rate of all other safety outcome measures at 1 year did not differ before and after implementation (***Table 2***).

**Figure 3.**
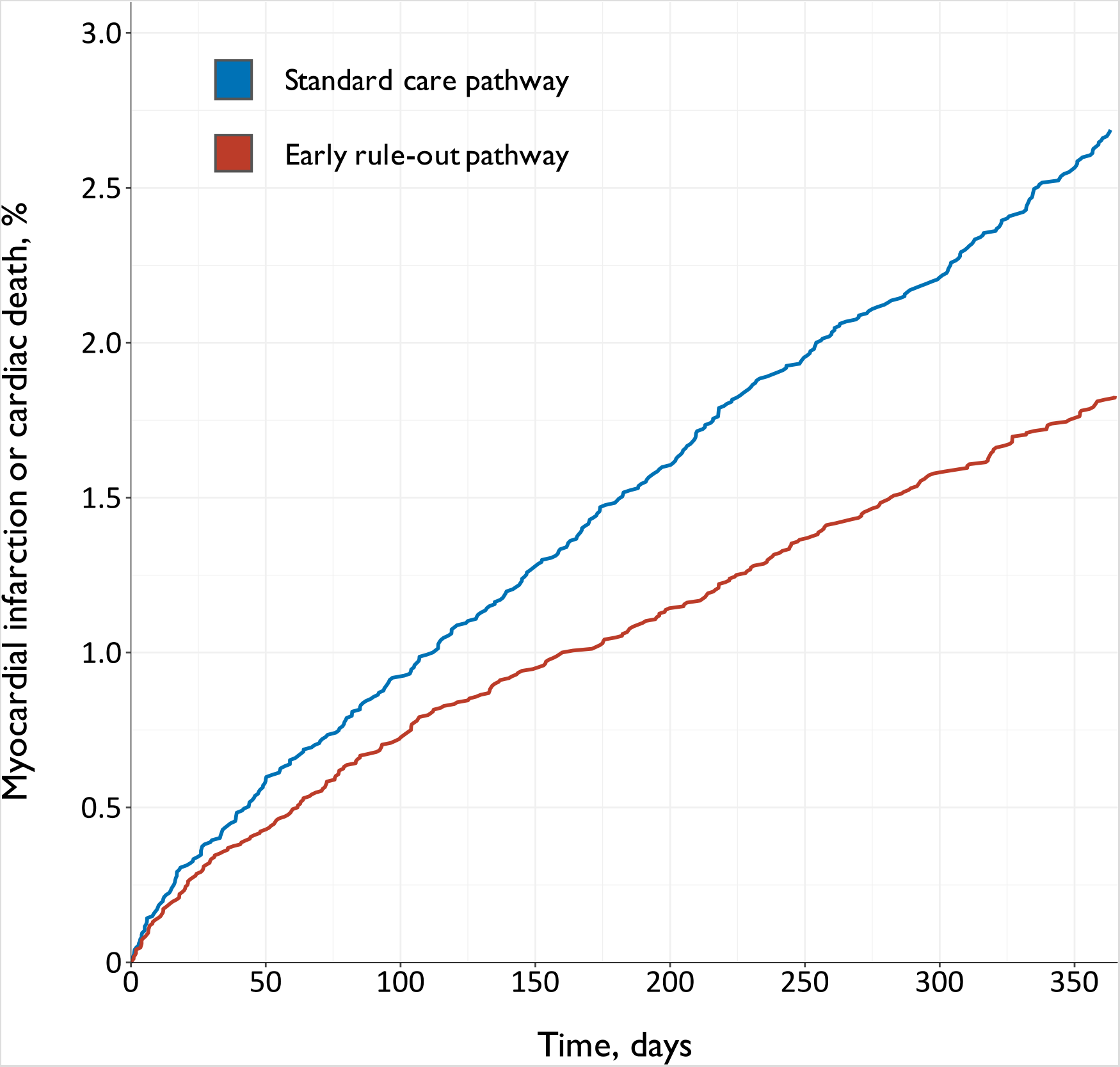
Myocardial infarction or cardiac death following discharge before and after implementation of the early rule-out pathway. Shown are cumulative incidence time-to-event curves for the primary safety outcome of myocardial infarction or cardiac death for patients evaluated before (blue line) and after (red line) implementation of the early rule-out pathway.

### Sensitivity Analysis in Calendar Matched Validation and Implementation Phases

In total 18,241 (58%) patients attended during the calendar-matched phases with 8,673 (48%) and 9,568 (52%) evaluated during the validation and implementation phase respectively. Length of stay was reduced from 10.6±4.1 to 6.8±4.0 hours (adjusted geometric mean ratio 0.65, 95% CI 0.62 to 0.68) before and after implementation of the early rule-out pathway. The primary safety outcome occurred in 43/8,673 (0.5%) and 23/9,568 (0.2%) patients at 30 days, with an adjusted odds ratio of 0.48 (95% CI 0.29 to 0.80, P = 0.005). The upper limit of our one-sided 95% confidence interval for the adjusted risk difference was −0.13%, which was below our superiority margin of 0%. The secondary safety outcome occured in 251/8,673 (2.9%) and 161/9,568 (1.7%) patients at 1 year (adjusted odds ratio 0.58, 95% CI 0.47 to 0.71, P< 0.001).

## Discussion

We evaluated the efficacy and safety of implementing an early rule-out pathway in 31,492 consecutive patients with suspected acute coronary syndrome. Introducing the pathway into clinical practice reduced length of stay by 3.3 hours and increased the proportion of patients avoiding hospital admission by 59%. Non-inferiority was not formally demonstrated, but the observed differences in myocardial infarction or cardiac death following discharge favoured the early rule-out pathway.

There are several strengths of our pragmatic trial design. First, we embedded our screening tool into the patient record to ensure we prospectively enrolled consecutive patients whom the attending clinician suspected acute coronary syndrome. This minimised the risk of selection bias, ensuring we did not limit our findings to low-risk patients or those presenting within working hours. Second, as the intervention was implemented at the hospital level, we did not seek individual patient consent. This reduced the risk of a Hawthorne effect where effectiveness is exaggerated through direct observation of clinical care by researchers. Third, our trial population was larger than the combined number of patients enrolled in 30 previous observational studies._26_,_27_ This ensured we had a greater number of events to evaluate safety. Finally, we combined hospital-level data with established registries to ensure follow up was complete in 99.8% of participants, and that our panel was able to adjudicate all safety outcome events.

The High-STEACS early rule-out pathway determines whether a patient with suspected acute coronary syndrome requires hospital admission or can be safely discharged. It is based on three principles. First, patients with very low troponin concentrations are at low-risk of cardiac events._6_ We defined the optimal risk stratification threshold as the highest concentration that gave a negative predictive value of > 99.5% for myocardial infarction or cardiac death at 30 days,_8_,_26_ to maximise the effectiveness of this approach whilst maintaining safety. Second, increasing concentrations above this risk stratification threshold on repeat testing may be important, even if they remain within the normal reference range, and these patients require admission to measure peak troponin concentration._18_ We define this using a change in concentration of ≥3 ng/L, based on the lowest measurable change that exceeds biological and analytical variation._28_ Third, to ensure our pathway is consistent with international guidelines,_4_ we applied the sex-specific 99_th_ centile as the threshold to identify patients who require hospital admission. Adherence was good across all seven acute hospitals which is testament to the simplicity of the pathway and should encourage adoption.

Whilst many pathways have been developed and validated that incorporate separate risk stratification and diagnostic thresholds,_12_,_29_ this is the first time that implementation has been evaluated in a prospective randomised controlled trial of consecutive patients. Here, we report substantial reductions in length of stay and increases in the proportion of patients avoiding hospital admission. Were these gains to be realised across healthcare systems, the benefits for both patients and providers would be substantial. In the US alone, more than 20 million patients with suspected acute coronary syndrome attend Emergency Departments each year._1_ A reduction in the length of stay of 3 hours could save more than $3.6 billion per annum on bed occupancy alone._30_ Despite these important reductions in length of stay, during the implementation phase the median stay was 6.8 hours, which is longer than reported in other evaluations of the implementation of early rule-out pathways._14_,_15_ This difference likely reflects our enrollment of all consecutive patients rather than selected patients who are less likely to have co-morbid conditions requiring hospital admission.

Implementation of our early rule-out pathway did not increase the rate of subsequent myocardial infarction or cardiac death. However, our results were highly sensitive to the model specification. Although non-inferiority was not concluded for the primary safety outcome at 30 days, in our prespecified sensitivity analysis restricted to calendar matched periods, the early rule-out pathway was superior to standard care at 30 days and 1 year. These divergent results may be due to the low event rate at 30 days and narrow randomisation phase leading to overfitting of the primary analysis model, additional secular changes not accounted for in the sensitivity analysis, or a true exposure-time effect whereby outcomes improved as the intervention became more firmly embedded into practice.

Is it plausible that the introduction of an early rule-out pathway could reduce the risk of subsequent cardiac events? By using a threshold well below the 99_th_ centile to risk stratify patients and by recognising that small changes in troponin concentration within the reference range may be important, we may have improved the evaluation of risk compared to using a single higher threshold to rule in and rule out myocardial infarction. This is supported by recent observational studies, which report that using the 99_th_ centile to rule out myocardial infarction at presentation and at 3 hours, misses 1 in 50 patients who would have been identified on serial testing 6-12 hours following the onset of symptoms._18_,_31_,_32_ Furthermore, our pathway encourages serial testing to safely rule out myocardial infarction in early presenters, which is now recognised by international guidelines._33_,_34_ More prospective trials in which clinical decisions are guided by new diagnostic approaches are needed to ensure our guidelines are based on the highest quality evidence.

Our findings add to those from a recently published randomised trial, which compared a 1-hour and 3-hour rule-out pathway._15_ In 3,378 patients the 1-hour pathway reduced length of stay by 60 minutes and increased discharge rates from 32% to 45%. The trial concluded non-inferiority for an endpoint of all-cause mortality or myocardial infarction within 30 days, although there was an increase in secondary safety outcome events in the 1-hour pathway arm. Due to a perceived lack of equipoise the monitoring committee recommended the trial stop recruitment with just two-thirds of the target population enrolled, and only one patient had a type 1 myocardial infarction following discharge in each arm.

We acknowledge several potential study limitations. Whilst the early rule-out pathway was implemented across three steps in the randomisation phase, we had to accept flexibility in the date of implementation (***Supplement***). This limited our ability to interpret a planned sensitivity analysis within the randomisation phase, when there were sites using both the standard care and early rule-out pathway. Additionally, we enrolled fewer than the 38,994 patients anticipated in our sample size calculations, and identified fewer safety outcome events at 30 days. We believe this in part contributed to modelling issues when attempting to evaluate the safety outcome at 30 days. However, more than 700 patients had a myocardial infarction or cardiac death at 1 year, and the rates of all secondary outcome measures were lower following implementation of the early rule-out pathway. Our pathway has been validated for use with two troponin I assays and a troponin T assay,_17_,_18_,_35_ and whilst it is likely to perform similarly for all high-sensitivity assays, further research is required to confirm this.

In conclusion, implementation of an early rule-out pathway for myocardial infarction substantially reduced length of stay and increased the proportion of patients avoiding hospital admission with no increase in adverse cardiac events. Adoption of this approach would have major benefits for both patients and healthcare providers.

## Data Availability

The HiSTORIC trial makes use of multiple routine electronic health care data sources that are linked, deidentified and held in our national safe haven, which is accessible by approved individuals who have undertaken the necessary governance training. Summary data can be made available upon request to the corresponding author.

## Author Contribution and Transparency Statement

The HiSTORIC Investigators contributed to the conception or design of the work, or the acquisition, analysis, or interpretation of data for the work. They were involved in drafting the manuscript and revising it, and have given final approval of the version to be published. The study sponsor had no role in the study design, collection of data, analysis and interpretation of data, writing of this report or decision to submit for publication. The corresponding author affirms that the manuscript is an honest, accurate, and transparent account of the study being reported and that no important aspects of the study have been omitted.

## Acknowledgements

This trial was funded by the British Heart Foundation (BHF) (PG/15/51/31596) with support from BHF Research Excellence Awards (RE/18/5/34216; RE/18/6134217). AA, KL and ARC are supported by a Clinical Lectureship from the Chief Scientist Office (PCL/18/05), a Clinical Research Training Fellowship from the BHF (FS/18/25/33454), and a Clinical Lectureship from the Scottish Clinical Research Excellence Development Scheme, respectively. DEN, ASVS, and NLM are supported by the BHF through the award of a Chair (CH/09/002), an Intermediate Clinical Research Fellowship (FS/19/17/34172), and the Butler Senior Clinical Research Fellowship (FS/16/14/32023), respectively. PDA is supported by a National Heart Foundation of New Zealand Senior Fellowship (1844). DEN is a recipient of a Wellcome Trust Senior Investigator Award (WT103782AIA). CJW and RP were supported by NHS Lothian through the Edinburgh Clinical Trials Unit. The funders played no role in the design, conduct, data collection, analysis or reporting of the trial. We would like to thank researchers from the Emergency Medicine Research Group Edinburgh and the Edinburgh Clinical Trials Unit for their support during the conduct of this trial.

